# Mathematical assessment of wastewater-based epidemiology to predict SARS-CoV-2 cases and hospitalizations in Miami-Dade County

**DOI:** 10.1101/2024.04.15.24305858

**Authors:** Binod Pant, Salman Safdar, Calistus N. Ngonghala, Abba B. Gumel

## Abstract

This study presents a wastewater-based mathematical model for assessing the transmission dynamics of the SARS-CoV-2 pandemic in Miami-Dade County, Florida. The model, which takes the form of a deterministic system of nonlinear differential equations, monitors the temporal dynamics of the disease, as well as changes in viral RNA concentration in the county’s wastewater system (which consists of three sewage treatment plants). The model was calibrated using the wastewater data during the third wave of the SARS-CoV-2 pandemic in Miami-Dade (specifically, the time period from July 3, 2021 to October 9, 2021). The calibrated model was used to predict SARS-CoV-2 case and hospitalization trends in the county during the aforementioned time period, showing a strong correlation (with a correlation coefficient *r* = 0.99) between the observed (detected) weekly case data and the corresponding weekly data predicted by the calibrated model. The model’s prediction of the week when maximum number of SARS-CoV-2 cases will be recorded in the county during the simulation period precisely matches the time when the maximum observed/reported cases were recorded (which was August 14, 2021). Furthermore, the model’s projection of the maximum number of cases for the week of August 14, 2021 is about 15 times higher than the maximum observed weekly case count for the county on that day (i.e., the maximum case count estimated by the model was 15 times higher than the actual/observed count for confirmed cases). This result is consistent with the result of numerous SARS-CoV-2 modeling studies (including other wastewater-based modeling, as well as statistical models) in the literature. Furthermore, the model accurately predicts a one-week lag between the peak in weekly COVID-19 case and hospitalization data during the time period of the study in Miami-Dade, with the model-predicted hospitalizations peaking on August 21, 2021. Detailed time-varying global sensitivity analysis was carried out to determine the parameters (wastewater-based, epidemiological and biological) that have the most influence on the chosen response function - the cumulative viral load in the wastewater. This analysis revealed that the transmission rate of infectious individuals, shedding rate of infectious individuals, recovery rate of infectious individuals, average fecal load *per* person *per* unit time and the proportion of shed viral RNA that is not lost in sewage before measurement at the wastewater treatment plant were most influential to the response function during the entire time period of the study. This study shows, conclusively, that wastewater surveillance data can be a very powerful indicator for measuring (i.e., providing early-warning signal and current burden) and predicting the future trajectory and burden (e.g., number of cases and hospitalizations) of emerging and re-emerging infectious diseases, such as SARS-CoV-2, in a community.

## 1 Introduction

The use of wastewater to study the spread of infectious disease (based on isolating pathogens from the stool of some individuals infected with the disease) has a long history, dating back to the pioneering work of John Snow (considered the *“Founding Father”* of modern epidemiology) in 1854, which showed that cholera-contaminated water from a Broad Street pump was the source of the cholera transmission in the Soho district of London [1, 2]. In the late 1930s and early 1940s, a Yale Poliomyelitis Study Unit led by John Paul and James Trask detected *poliomyelitis* (polio) virus in sewage collected in multiple cities in the United States with active polio epidemics, including Charleston, South Carolina; Detroit, Michigan; and Buffalo, New York, by injecting wastewater in the endemic regions into monkeys and observing to see if the monkeys acquire the polio infection [3–5]. Their experiment showed that wastewater could potentially be used as an indicator of disease activity in a community. However, their method involving monkeys was costly, slow, and inefficient as the monkeys died from other chemicals or pathogens that were present in the wastewater injected into them [6]. In the modern times, the availability of efficient and safe methods for detecting viral RNA in wastewater (such as *reverse transcription–polymerase chain reaction* (RT-PCR)), wastewater-based surveillance of disease activity has become an effective and resource-efficient tool for gathering crucial/important community-level public health information during outbreaks of infectious diseases [7, 8]. For instance, the United States Centers for Disease Control and Prevention (CDC) launched the National Wastewater Surveillance System (NWSS) in September of 2020 to “coordinate and build the nation’s capacity to track the presence of SARS-CoV-2, the virus that causes COVID-19, in wastewater samples collected across the country” [9].

Wastewater-based surveillance epidemiology has numerous benefits. It, for instance, is a less invasive disease surveillance mechanism, in comparison to conducting mass testing to detect the level of spread of a pathogen in the community [10]. Furthermore, in the context of COVID-19, while a kit for rapid antigen tests can cost as little as 2 dollars to manufacture, bidding wars between health systems, state governments and employers can contribute to much higher prices [11]. For example, during the year 2021, the state of South Carolina was reported to have paid as high as 130 for some of its rapid test kits [11]. Ngwira *et al*. [12] estimated the economic costs of conducting wastewater-based environmental surveillance for SARS-CoV-2 in Blantyre, Malawi and Kathmandu, Nepal, estimating monthly costs of approximately $6,175 to $8,272 for Blantyre and $16,756 to $30,050 for Kathmandu, respectively. It is further estimated that the cost *per* person in the catchment area annually ranged from $0.07 to $0.10 in Blantyre and $0.07 to $0.13 in Kathmandu [12], respectively. Wastewater surveillance, when combined with other forms of public health surveillance and interventions, can result in a very effective control and mitigation of infectious diseases spreading in communities [13, 14]. For instance, during the SARS-CoV-2 pandemic, in 2020, researchers from the University of Arizona, Tucson, used wastewater samples from campus dorms to identify which dorms were infected (i.e., wastewater from which student dormitory contained viral RNA), subsequently leading to mass testing of students in the infected dorm (and the detection and quarantine of two asymptomatic cases) [15, 16]. It has also been suggested that wastewater could potentially act as a “leading indicator” or an “early warning signal” for a disease outbreak [17–21]. Olesen *et al*. [18] state that, “the biological principle behind wastewater as a leading indicator is that many infected individuals shed the virus in stool before they develop symptoms and thus also before they seek medical care”. Finally, wastewater genomic surveillance can provide insight into disease variants (or strains) co-circulating in the community [22, 23]. Wastewater surveillance in airports and on aircraft has been proposed as a low-cost mechanism to monitor SARS-CoV-2 variants entering a region [24–26]. All of these reasons not only highlight the importance of using wastewater surveillance to detect or mitigate disease activity, but also hint at the high utility of incorporating wastewater data into mathematical models for the spread of emerging and re-emerging infectious diseases in a community to enhance their prediction capacity.

Numerous mathematical models that use wastewater surveillance data have been developed and used to study the transmission dynamics and control of numerous emerging and re-emerging infectious diseases in communities. Before specifically discussing some of the COVID-19 mathematical models that used wastewater data, it is intuitive to highlight the fact that numerous models for cholera epidemics have already accounted for the concentration of the bacterium that causes the disease (*V. cholerae*) in water [27–32]. The key difference between the COVID-19 wastewater models to be discussed and the cholera models is that, while in the latter contaminated water is a source of infection, in the former, the data collected from the wastewater (sewage) is used as an indicator of the level of spread of the disease in the community. The state of Israel was certified as polio-free by the World Health Organization in June 2002 [33]. About 11 years later, wild-type *poliovirus 1* (WPV1) was detected during routine sewage samples collected between the 7th to the 13th of April 2013 [33, 34]. Oral polio vaccine (OPV), a weakened poliovirus given in the form of oral drops [35], was discontinued and replaced by inactivated poliovirus vaccine (IPV) only policy in Israel in 2005 [34, 36]. This change in policy was due to the risk of the emergence of circulating vaccine-derived poliovirus (cVDPV) and vaccine-associated paralytic poliomyelitis [36]. Although both types of the polio vaccine provide excellent protection against the disease, only OPV is able to prevent transmission (by inducing “strong gut immunity that blocks transmission of the virus, which is shed in the stool and spread largely through fecal-oral contamination” [36]). In response to the sudden reappearance of polio, in early August 2013, Israel launched a supplementary OPV vaccination program, by September 20, 2023 and 60% of the targeted Israeli children had been vaccinated with bivalent OPV [34]. The last two positive samples detected in Israel were on October 2013 and February 2014 [37], thus effectively ending the outbreak in less than a year’s time. The silent spread of polio in Israel in 2013-2014 and its subsequent suppression before the detection/development of a single paralytic polio case highlights the importance of sewage-based surveillance.

In the context of the COVID-19 pandemic, several studies have shown a correlation between SARS-CoV-2 RNA levels in wastewater and COVID-19 cases in the community [38–41]. Statistical models have used wastewater data to predict COVID-19 cases [42], hospital admission [43–46] and the effective reproduction number [47]. Furthermore, compartmental models that incorporate wastewater data have also been used to achieve the same objective. For instance, McMahan *et al*. [48] used an *SEIR* model and wastewater surveillance data to show that the number of model-predicted COVID-19 cases in Clemson University, South Carolina, between the time period from May 27, 2020 to August 25, 2020, was 11 (95% CI 4.2–17.5) times higher than the confirmed (i.e., observed) cases. Phan *et al*. [49] proposed an SEIR-V (where *V* is the viral load in wastewater) model that uses wastewater data to also show that the model-predicted COVID-19 cases in the Greater Boston area, during the period from October 2, 2020 to January 25, 2021, were 8.3 to 10.2-fold higher than confirmed cases in Greater Boston area from October 2, 2020 to January 25, 2021. Nourbakhsh *et al*. [50] extended the *SEIR* model to explicitly account for various classes of infectious individuals (symptomatic, asymptomatic and hospitalized) and recovered individuals who are still shedding the virus in feces. The authors also accounted for the delay and degradation of RNA from shedding to sampling.

This current study is based on using a relatively simple SEIR epidemiology-wastewater model to predict the burden of the SARS-CoV-2 and disease-related hospitalizations in a population. Specifically, the model will be used to predict the case and hospitalization trends for Miami-Dade County, Florida. Although various compartmental models have been used to predict the burden of COVID-19 at a county and state level using SARS-CoV-2 case and mortality [51–53] and case data [54, 55], the utility of these models is affected by decisions to halt the collection of such data, as was the case with SARS-CoV-2 (when the Federal COVID-19 Public Health Emergency (PHE) Declaration expired on May 11, 2023; and the CDC’s authorization to collect certain types of public health data (most notably COVID-19 cases) also expired) [56, 57]. Furthermore, the confirmed case data reported by the CDC does not include asymptomatic COVID-19 cases (i.e., the confirmed case count does not accurately reflect the true burden of the disease) [58]. Additionally, the widespread availability of free at-home COVID-19 test kits since January 2022 [59] impacted the accuracy of the official testing count (due to under-reporting of positive at-home test results) [60]. These factors, taken in totality, suggest that compartmental models for the spread and control of the SARS-CoV-2 used after the halting of data collection in May 2023 may fail to capture the correct dynamics of the disease

Finally, the number of SARS-CoV-2-related deaths occurring at a county level may be low enough (or may occur infrequently enough) that fitting an ODE model to daily or weekly mortality data may pose additional challenges. This also potentially limits the utility of using mortality data (after May 2023) to fit compartmental models for the SARS-CoV-2 pandemic. Wastewater surveillance at the county level provides a cost-effective alternative to remedy the unavailability of reliable epidemiological (case and mortality) data for use to predict the transmission dynamics and control of the disease at the county level. The use of wastewater surveillance data (as an indicator for disease activity) is the main focus of this study. The paper is organized as follows. The wastewater-based epidemiology (WBE) model will be formulated in Section 2. Specifically, while the equations describing the disease transmission dynamics in humans are described in Section 2.1, the equations for the dynamics of SARS-CoV-2 RNA concentration in the wastewater are described in Section 2.2. Asymptotic stability results for the associated disease-free equilibrium of the model are given in Section 2.3. The model is fitted, using weekly wastewater data for the county, in Section 3. Predictions for the number of weekly hospitalizations in the county are also made in this section. Detailed time-varying global sensitivity analysis is carried out in Section 4 to determine the parameters (wastewater-based, epidemiological and biological) that have the most influence on the chosen response function (namely, the cumulative viral load in the wastewater). Numerical simulations are carried out in Section 5. The main results of the study, along with its limitations, are discussed and summarized in Section 6.

## 2 Wastewater-based Epidemiology (WBE) Model

In order to design the SARS-CoV-2 transmission model that incorporates wastewater data, the total human population at time *t*, denoted by *N* (*t*), is split into mutually-exclusive compartments of susceptible (*S*(*t*)), exposed (i.e., newly infected but not infectious and not shedding virus into the wastewater system; *E*(*t*)), infectious and shedding viral RNA into the wastewater (*I*(*t*); this population includes both asymptomatic and symptomatic infectious individuals), hospitalized and shedding viral RNA into the wastewater (*H*(*t*)), recovered and shedding viral RNA into the wastewater (*J*(*t*)) and recovered but not shedding viral RNA into the wastewater (*R*(*t*)), individuals, so that:

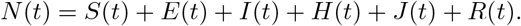

Let *V* (*t*) represent the concentration of viral RNA (shed through feces) in the wastewater at time *t*. Furthermore, let *W* (*t*) represents the cumulative amount of measured SARS-CoV-2 concentration in the wastewater at time *t*, after accounting for decay and delay of RNA from excretion to arrival, and other factors such as uncertainty in measurement. The equations for the rate of change of the various compartments are derived below.

### 2.1 Equations for SARS-CoV-2 dynamics between humans

Susceptible individuals acquire infection, following effective contact with infectious individuals (in the *I* and *H* classes), at a rate *λ*(*t*) (*force of infection*), given by:

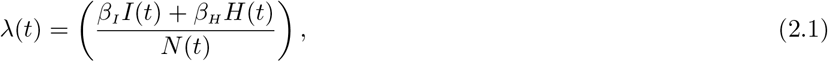

where *β*_*I*_ and *β*_*H*_ represent the effective contact rate of infectious individuals in the infectious (*I*) and hospitalized (*H*) compartments. Newly-infected individuals in the exposed class (*E*) progress to develop clinical symptoms of SARS-CoV-2 at a rate *σ*. Infectious individuals are hospitalized at a rate *ϕ*. Infectious and hospitalized individuals recover (while also shedding viral RNA into the wastewater) at a rate *γ*_*I*_ and *γ*_*H*_, respectively (i.e., *γ*_*I*_ and *γ*_*H*_, represent the rates at which individuals in the *I* and *H* classes progress to the *J* class, respectively, upon recovery from SARS-CoV-2 infection). Finally, recovered individuals who shed viral RNA into the wastewater (i.e., individuals in the *J* class) move to the no-viral-shedding recovered class (*R*) at a rate *γ*_*J*_. It follows, based on the above assumptions and derivations, that the wastewater-epidemiology (WBE) epidemic model for the transmission of SARS-CoV-2 in a community is given by the following deterministic system of nonlinear differential equations (where a dot represents differentiation with respect to time *t*):

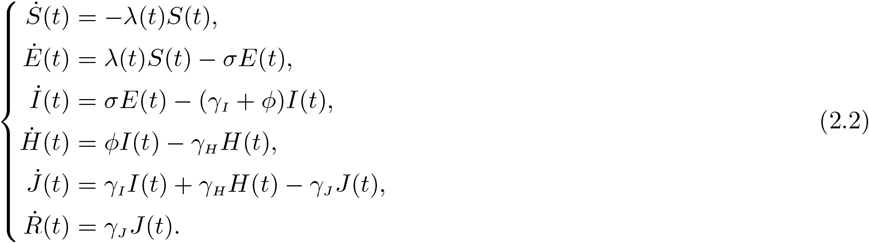

### 2.2 Equations for dynamics of viral concentration in the wastewater

To derive the equations for the dynamics of viral concentration in the wastewater system, we assume, first of all, that the quantity of viral shedding (through feces) into the wastewater at time *t*, denoted by *V* (*t*), increases by the viral shedding of individuals in the infectious (*I*), hospitalized (*H*) and recovered but shedding (*J*) classes. Let *αζ*_*I*_, *αζ*_*H*_ and *αζ*_*J*_ represent the rate at which individuals in the *I, H* and *J* compartments shed viral RNA into the wastewater system in the community. Here, *α* is the average fecal load (gram *per* unit time *per* person) and *ζ*_*k*_ (with *k* = {*I, H, J*}) is the average viral shedding rate of an individual in *k* compartment (RNA copies *per* gram) [49]. The quantity *V* is reduced at a rate *d*_*V*_ (where 1*/d*_*V*_ is the average time from the excretion of feces to the collection of a sample at the wastewater treatment plant). Furthermore, only a certain proportion of the viral RNA shed into the wastewater is measurable (denoted by a parameter *η*) due to a number of complex processes that affect RNA (for example, due to temperature-related decay) [61,62]. Hence, the cumulative amount of the viral RNA measured in (or collected from) the wastewater, denoted by *W* (*t*), is increased at a rate *ηd*_*V*_. Thus, based on the above assumptions and derivations, the equations for the rate of change of the viral concentrations *V* and *W* are given, respectively, by:

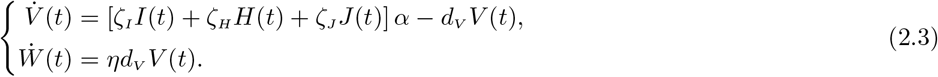

Let 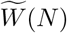 represent the cumulative measurement of viral RNA in wastewater at time *N*. Hence, 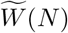 can be obtained from the integral:

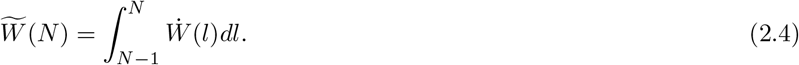

The WBE model, defined based on coupling the equations for the dynamics of SARS-CoV-2 in humans with those for the viral dynamics within the wastewater system, is given by the system of equations {(2.2),(2.3)}. Figure 1 depicts the flow diagram of the WBE model, and the state variables and parameters are described in Tables 1 and 2, respectively.

**Table 1:** Description of the state variables of the WBE model {(2.2),(2.3)}.

| State variable | Description |
| --- | --- |
| $S$ | Number of susceptible individuals |
| $E$ | Number of exposed (newly infected but not infectious or shedding) individuals |
| $I$ | Number of infectious (and shedding) individuals |
| $H$ | Number of hospitalized (and shedding) individuals |
| $J$ | Number of recovered (i.e., not infectious) individuals who are still shedding virus through feces |
| $R$ | Number of recovered (i.e., not infectious) individuals who are shedding virus through feces |
| $V$ | Quantity of viral RNA shed through feces |
| $W$ | Cumulative measurement of viral RNA in wastewater |

**Table 2:** Description of the parameters of the WBE model {(2.2),(2.3)}.

| Parameter | Description |
| --- | --- |
| $\beta_I(\beta_H)$ | Effective contact rate of individuals in the $I(H)$ compartment |
| $\sigma$ | Progression rate from exposed ($E$) to $I$ class |
| $\gamma_k$ ($k = \{I, H, J\}$) | Recovery rate for individuals in $k$ class |
| $\phi$ | Rate of at which individuals in $I$ class get hospitalized |
| $\alpha$ | Average fecal load *per person per unit time* |
| $\zeta_k$ ($k = \{I, H, J\}$) | Average viral shedding *per gram* from individuals in $k$ compartment |
| $\alpha\zeta_k$ ($k = \{I, H, J\}$) | Rate at which individuals in $k$ class shed |
| $1/d_V$ | Average time from excretion of feces to collection at the treatment plant |
| $\eta$ | Proportion of shed viral RNA that is not lost in sewage by measurement time |
| $\eta d_V$ | Progression rate of viral RNA from from $V$ to $W$ class |

**Figure 1:**
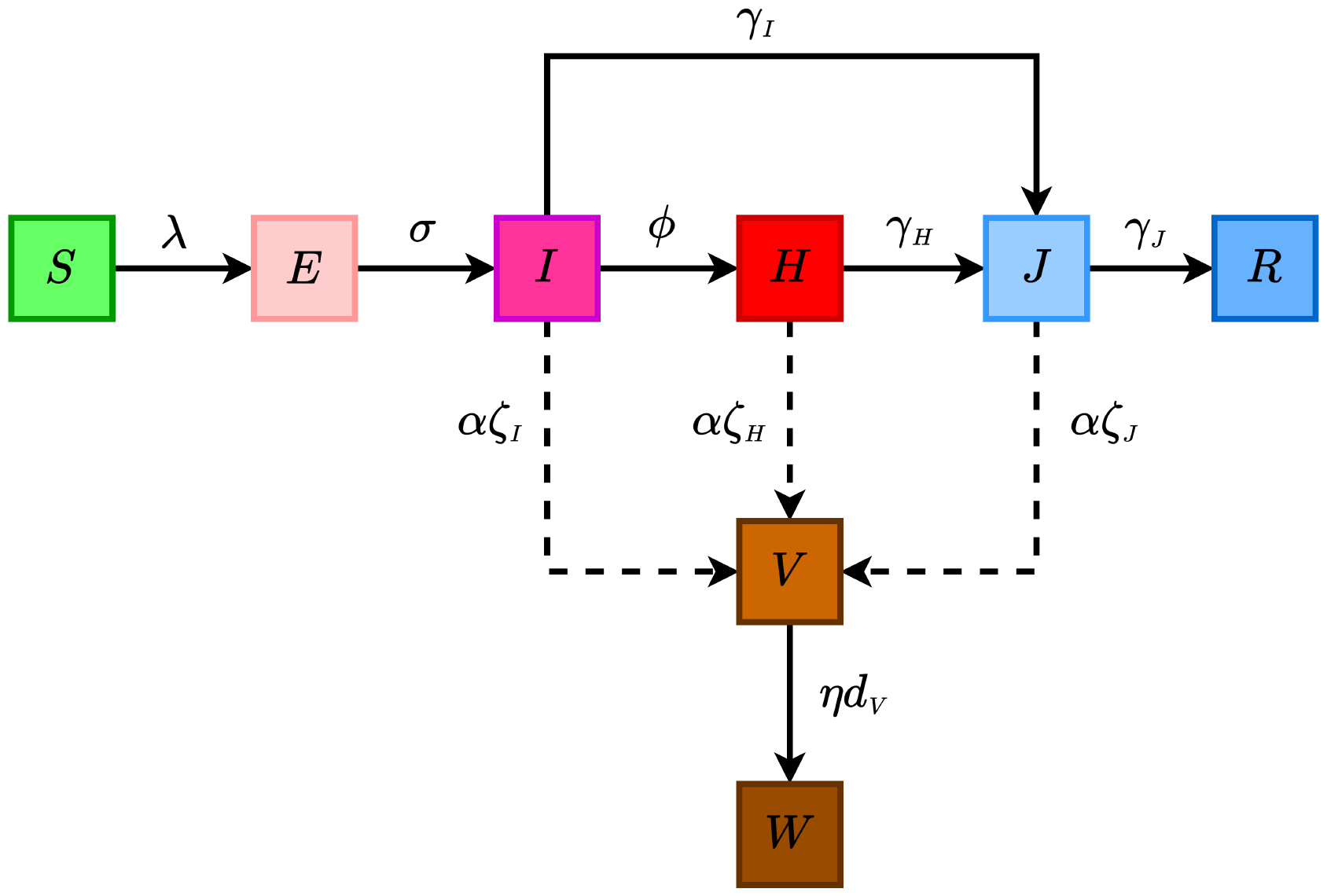
Flow diagram of the wastewater-based epidemiology (WBE) model {(2.2),(2.3)}, where *λ* is defined in Equation (2.1). The dashed lines represent the rate of fecal viral RNA shedding by individuals in the infectious (*I*), hospitalized (*H*) and recovered (*J*) compartments. The state variables and parameters of the model are described in Tables 1 and 2, respectively.

The main assumptions made in the formulation of the WBE model {(2.2),(2.3)} include:

a. closed population (i.e., no influx of people into or out of the population);
b. well-mixed homogeneous population (i.e., everyone is equally likely to mix with everyone else);
c. exponentially-distributed waiting times in each epidemiological compartment;
d. natural recovery induces permanent immunity against future infection (although there is evidence for reinfection with COVID-19 in some recovered individuals [63, 64])
e. the infectious compartment (*I*) consists of individuals who are both asymptomatically- and symptomatically-infectious. Although numerous SARS-CoV-2 modeling studies, such as those in [52, 65–68], have shown that asymptomatic individuals account for more new cases than symptomatic individuals, we lump the asymptomatic and symptomatic infectious individuals into one epidemiological compartment (*I*(*t*)) for mathematical tractability. This assumption is also necessitated due to the lack of reliable data, at the current moment, on the level of viral shedding by asymptomatic infectious individuals.

The WBE model (2.2),(2.3)} is an extension of various epidemiological models that incorporate wastewater dynamics [48,49]. In particular, it extended the model in [49] by, *inter alia*:

i. Adding compartment of (and viral shedding by) hospitalized individuals (hospitalized individuals are not explicitly accounted for in [49]).
ii. Adding the compartment *J* of recovered individuals who still shed viral RNA into the wastewater (this compartment was not accounted for in [49]).
iii. Using standard incidence to model the infection rate (mass action incidence was used in [49]).
iv. Modeling viral dynamics in the wastewater system using two equations, accounting for (a) the viral RNA shed by individuals in the community and (b) the viral RNA measured at the wastewater treatment plant (only one equation was used to model the wastewater component in [49]).

### 2.3 Asymptotic stability of disease-free equilibria

In this section, the local asymptotic stability of the disease-free solutions of the WBE model will be analysed to determine conditions, in parameter space, for the control or possible persistence of a SARS-CoV-2 outbreak in the community. It should, first of all, be noted that the sub-model describing the disease dynamics in humans (i.e.,Equation (2.2)) is decoupled from the equation for the wastewater dynamics (Equation (2.3)). Thus, the asymptotic dynamics of the WBE model reduces to the analysis of the disease-free equilibria of the sub-model (2.2).

It can be seen that the model (2.2) has a continuum of disease-free equilibria (DFE), given by [52, 65]:

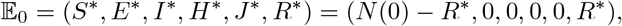

where *N* (0) is the initial total population, 0 < *S*^*\**^ ≤ *N* (0), 0 ≤ *R*^*\**^ ≤ *N* (0) and 0 < *S*^*\**^ + *R*^*\**^ ≤ *N* (0). It is convenient to define the total population at the DFE as *N*^*\**^ = *S*^*\**^ + *R*^*\**^. Since our epidemic model does not account for disease-induced mortality (i.e., the population is asymptotically constant), *N*^*\**^ = *N* (0). The asymptotic stability of the DFE of the model (2.2) can be analysed using the *next generation operator method* [69, 70]. *Using the notation in [69], it can be seen that the non-negative matrix of new infection terms (denoted by F*) and the M-Matrix of linear transition terms in the infected compartments of the model (denoted by *V*) are given, respectively, by:

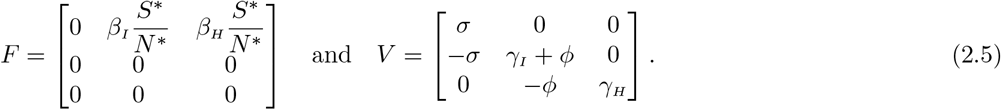

It is also convenient to define the quantity (where *ρ* is the spectral radius) [69, 70]:

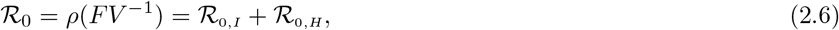

where,

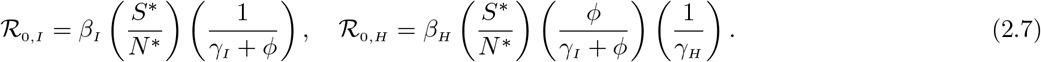

The quantity *ℛ*_0_ is the *basic reproduction number* of the model (2.2) (and, hence, of the WBE model {(2.2),(2.3)}). It is a measure of the average number of new cases generated by a typical infected individual if introduced into a population that is completely susceptible. The quantities *ℛ*_0,*I*_ and *ℛ*_0,*H*_ are the constituent basic reproduction numbers for the average number of new cases generated by infectious and hospitalized individuals, respectively. The result below follows from Theorem 2 of [69].

#### Theorem 2.1.

*The continuum of disease-free equilibria* (E_0_) *of the model* (2.2) *is locally-asymptomatically stable (LAS) if ℛ*_0_ < 1.

The epidemiological implication of Theorem 2.1 is that a small influx of COVID-19 cases will not generate a large outbreak in the community if *ℛ*_0_ < 1. In the case of epidemic models, such as the sub-model (2.2), the epidemiological requirement of having *ℛ*_0_ < 1, while sufficient, is not necessary for the elimination of the disease from the community [52, 65]. In other words, unlike in an endemic model (with demographic birth and death processes, guaranteeing an influx of new susceptible individuals into the population), the disease will die out (with time) under the assumptions of an epidemic model regardless of the value of the reproduction number (it should be noted that models for new epidemics, such as (2.2), always assume a large, closed population, with no influx of people into or out of the population, in addition to the assumption that the timescale of the epidemic is smaller than the demographic timescale [71]. Consequently, the susceptible population can decrease as more people acquire the infection but is not replenished, thus eventually leading to the elimination of the disease even if the reproduction number exceeds unity.

#### Remark 2.1.

*In the case that ℛ*_0_ > 1, *the epidemic will grow to a peak and then eventually decline to zero [52, 65, 72]*.

## 3 Data-fitting and Parameter Estimation

The wastewater-based epidemiology (WBE) model {(2.2),(2.3)} has 13 parameters. While the values of 8 parameters is known from literature, the values of the remaining 5 parameters (namely the the parameters for the effective contact rate of infectious (*β*_*I*_) and hospitalized (*β*_*H*_) individuals, the rate of hospitalization (*ϕ*), the average fecal load *per* person (*α*) and the proportion of shed viral RNA that arrives at the treatment plant (*η*) will be obtained by fitting the model with the wastewater data from the community. Specifically, the WBE model {(2.2),(2.3)} will be fitted using the weekly-measured wastewater data (obtained from Biobot Analytics [73]) for the COVID-19 pandemic in the Miami-Dade county, Florida, for the period from July 3, 2021 to October 9, 2021. Additionally, some of the initial values of the state variables of the model (namely, *E*(0), *I*(0), *J*(0) and *R*(0)) will be fitted from the data as well (see the details in the caption of Figure 3). Miami-Dade county has three wastewater treatment plants (WWTPs): the north district WWTP, the central district WWTP and south district WWTP, servicing approximately 780,000, 830,000 and 920,000 people, respectively [74]. U.S. Census Bureau estimates the total population of Miami-Dade county for July 1, 2022 to be 2,673,837 [75]. Hence, these wastewater treatment plants serve about 95% of the total population of Miami-Dade county. The map of the service area corresponding to each of the three treatment plants is given in Figure 2. The wastewater data to be used to fit the WBE model is processed by the company Biobot Analytics. Since the company provides average weekly viral RNA data collected from each of the three treatment plants in the county [73], we multiply the data by seven to convert it to total weekly viral RNA data (to align with the total weekly hospitalization data we are generating from the WBE model). The total weekly wastewater data is tabulated in Table B.1 of Appendix A). Finally, following [49], the total weekly viral load in sewershed is obtained by multiplying the weekly wastewater data by the total influent flow rate *per* week (estimated to be 300 million gallons *per* day or 2,100 million gallons *per* week for Miami-Dade county [76, 77]). It should be stated that the multiplication of the processed wastewater data (measured in copies/mL; see column 2 of Table B.1) by the influent flow rate (with unit mL/week) results in the weekly amount of RNA in wastewater (measured in copies/week).

**Figure 2:**
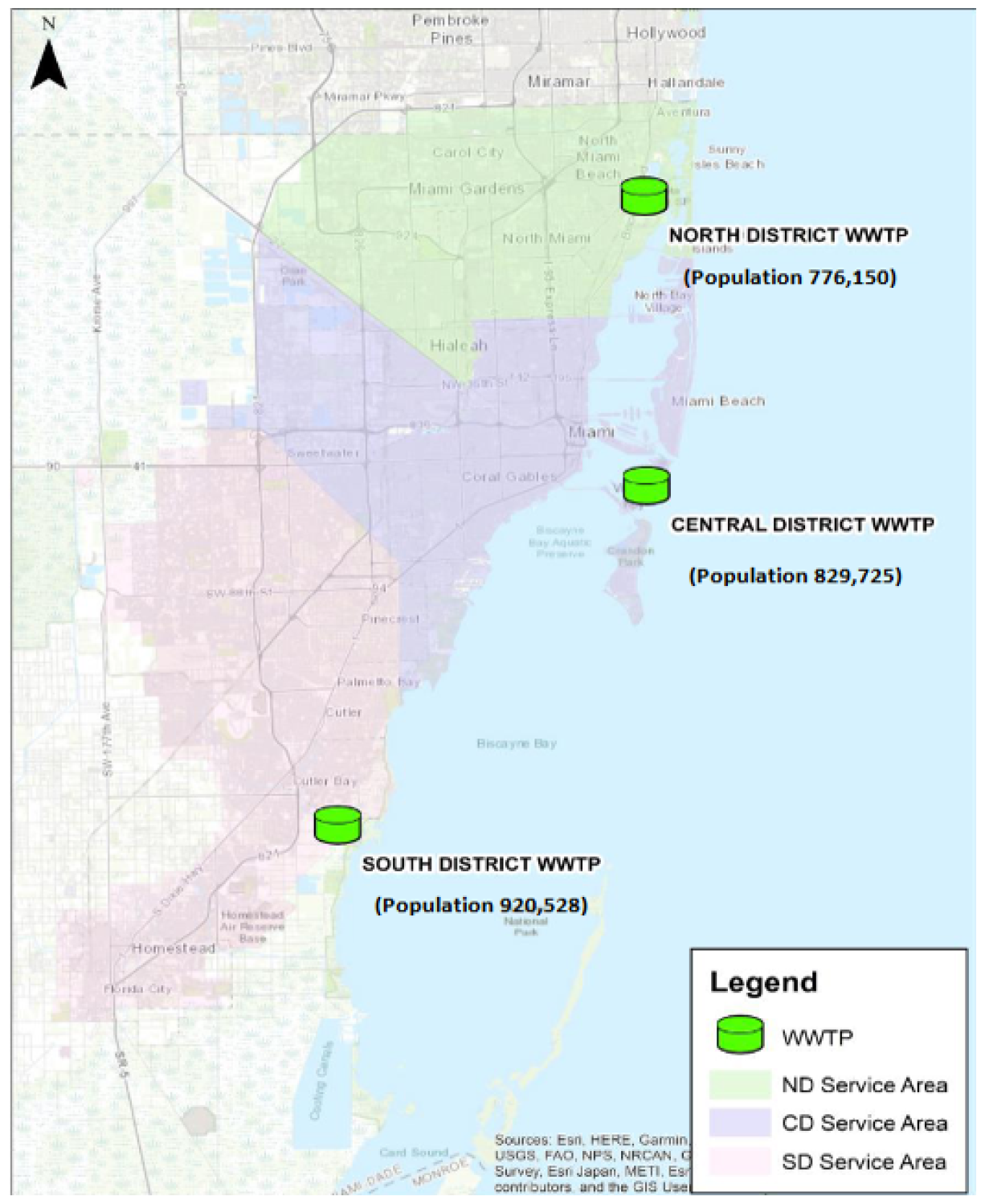
Map showing the three wastewater treatment plants (WWTP) in Miami-Dade county in Florida and their service area. The three treatment plants, namely the north district treatment plant, the central district treatment plant and the south district treatment plant, are marked in the map with green cylinder icons and their service area is depicted with green, violet and red shaded color in the map, respectively. The north, central and south district treatment plants service approximately 0.78, 0.83 and 0.92 million individuals, respectively. Source: Public Health Dashboard Report of January 30, 2023 by Miami-Dade County [74].

**Figure 3:**
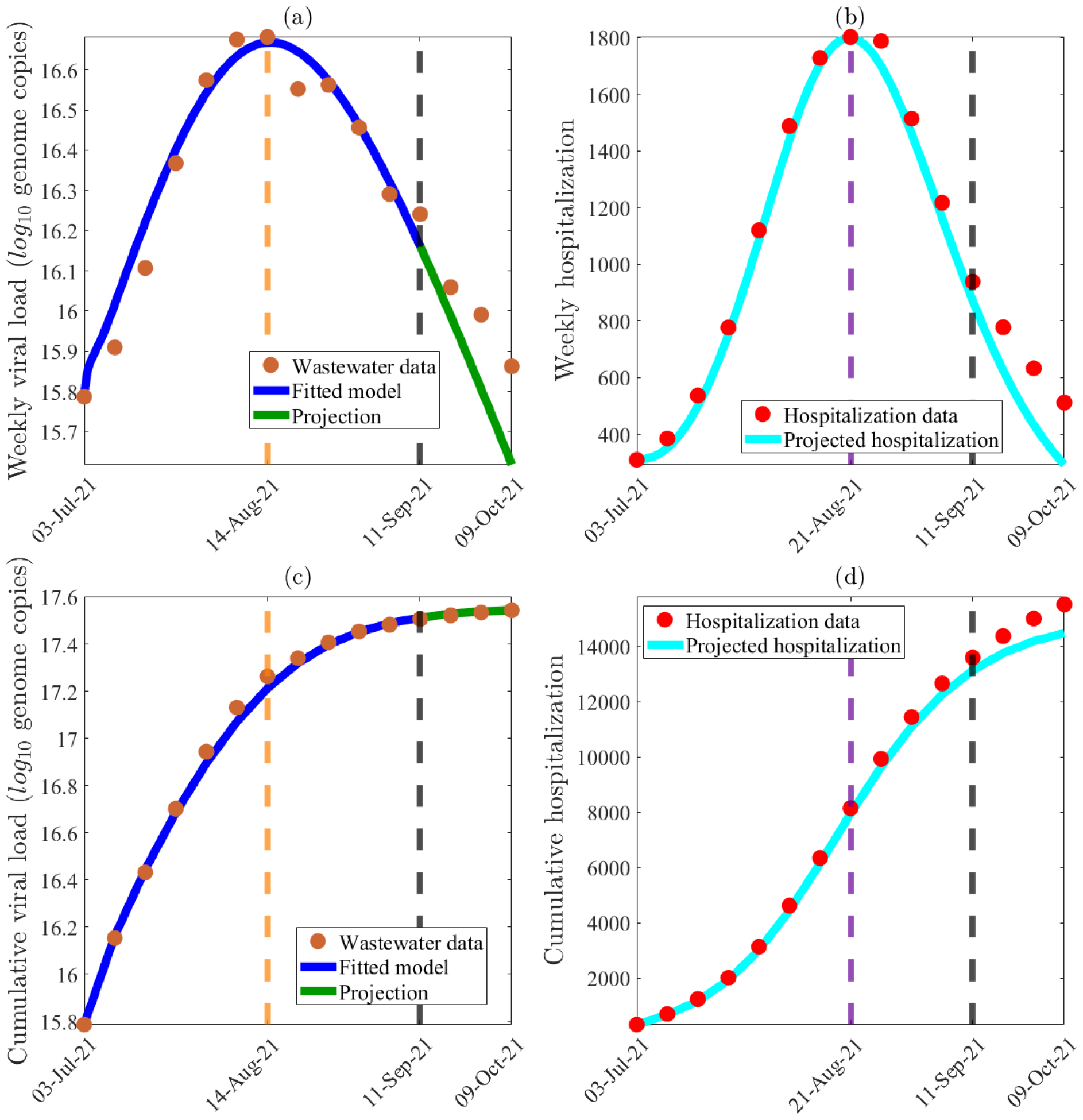
Data fitting, parameter estimation and projection of the model {(2.2), (2.3)} using weekly wastewater data for Miami-Dade County, Florida from July 3rd, 2021 to October 9th, 2021. (a) Least-squares fitting of the WBE model {(2.2), (2.3)}, showing weekly viral load in wastewater generated from the model, compared to the observed wastewater data (obtained from Biobot Analytics [73] and Sewer Department of Miami-Dade County [76]). (b) Simulation results of the WBE model {(2.2), (2.3)}, depicting the weekly COVID-19 hospitalization for Miami-Dade County, as a function of time, predicted by the WBE model using the fixed and estimated parameters tabulated in Tables 3 and 4, respectively, in comparison to the observed weekly hospitalization data for Miami-Dade County from July 3rd, 2021 to October 9th, 2021 (obtained from The New York Times [84]). (c) The model projected cumulative viral load in wastewater (depicted with a blue curve) is compared with the observed cumulative wastewater data for Miami-Dade County from July 3rd, 2021 to October 9th, 2021 (depicted with brown dots). (d) The model projected cumulative COVID-19 hospitalization (depicted with the cyan curve) is compared to the observed cumulative hospitalization data (depicted with red dots) for Miami-Dade County from July 3rd, 2021 to October 9th, 2021. The WBE model was fitted to the weekly wastewater data from July 1st, 2021 until September 11th, 2021 (depicted by a dashed black line in the figure), and the projection of the model output is compared with the observed wastewater data till October 9, 2021 (the projection period is depicted with a green curve). The vertical dashed orange and violet lines in Figures (a) and (b) respectively correspond to the time when weekly wastewater and hospitalization data peaks (i.e., August 14, 2021, and August 21, 2021, respectively). Four of the initial conditions of the WBE model (namely *E*(0), *I*(0), *J* (0) and *R*(0)) were estimated by fitting the model with the weekly wastewater data. The estimated values were *E*(0) = 17, 377, *I*(0) = 55, 592, *J* (0) = 3, 058 and *R*(0) = 2, 077, respectively. The initial condition for the number of hospitalized individuals (i.e., *H*(0)) is obtained from The New York Times to be 310 [84]. Using the U.S. Census data, the total initial population of Miami-Dade County is assumed to be *N* (0) = 2, 716, 940 [92,93]. Hence, the initial susceptible population is obtained as *S*(0) = *N* (0) *−* (*E*(0) + *I*(0) + *H*(0) + *J* (0) + *R*(0)). The remaining initial conditions are *V* (0) = 10^14.96^ (i.e., log_10_(*V* (0)) = 14.96) and *W* (0) = 10^15.79^ (i.e., log_10_(*V* (0)) = 15.79).

Before describing the data fitting of the WBE model {(2.2),(2.3)}, the values of the 8 known parameters (also known as *fixed parameters*) of the model are described below.

**Table 3:** Values of the fixed parameters of the WBE model {(2.2),(2.3)}.

| Parameter | Value | Source |
| --- | --- | --- |
| $\sigma$ | $7/3 \text{ week}^{-1}$ | [78] |
| $\gamma_I$ | $7/8 \text{ week}^{-1}$ | [79] |
| $\gamma_H$ | $7/8 \text{ week}^{-1}$ | [80] |
| $\gamma_J$ | $7/24 \text{ week}^{-1}$ | [50, 90, 91] |
| $\zeta_I$ | $4.49 \times 10^7 \text{ viral RNA per gram}$ | [49] |
| $\zeta_H$ | $4.49 \times 10^7 \text{ viral RNA per gram}$ | Assumed |
| $\zeta_J$ | $3.17 \times 10^4 \text{ viral RNA per gram}$ | Calculated (see (3.2)) |
| $d_V$ | $7 \text{ week}^{-1}$ | Assumed |

**Table 4:** Values of the fitted (estimated) parameters of the WBE model {(2.2),(2.3)}.

| Parameter | Estimated (fitted) Value |
| --- | --- |
| $\beta_I$ | $1.77 \text{ week}^{-1}$ |
| $\beta_H$ | $8.47 \times 10^{-3} \text{ week}^{-1}$ |
| $\phi$ | $5.12 \times 10^{-3} \text{ week}^{-1}$ |
| $\eta$ | 0.96 (dimensionless) |
| $\alpha$ | $3.28 \times 10^4 \text{ grams week}^{-1}$ |

### 3.1 Values of the fixed (known) parameters of the WBE model

The values of the 8 known parameters of the WBE model {(2.2),(2.3)} (tabulated in Table 3) are described as follows: the incubation period for COVID-19 is estimated to be 3 days [78], so we set the transition parameter (*σ*) from the exposed to the infectious class to be *σ* = 1*/*3 *per* day or 7/3 *per* week. The recovery time for infectious individuals (1*/γ*_*I*_) and hospitalized individuals (1*/γ*_*H*_) is 8 days [79, 80]. Thus, *γ*_*I*_ = *γ*_*H*_ = 1*/*8 *per* day or 7/8 *per* week. Upon recovery, newly-recovered individuals (i.e., individuals in the *J* compartment) continue to shed viral RNA into the wastewater for 24 days and cease to shed afterwards, transitioning to the *R* class). Hence, *γ*_*J*_ = 1*/*24 *per* day or 7/24 *per* week. Phan *et al*. [49] fitted viral shedding in SARS-CoV-2 hospitalized patients’ stool data in [79] (given in log_10_ viral RNA copies *per* gram) to obtain a function, *f* (*t*), that describes the temporal fecal viral shedding profile, and is given by:

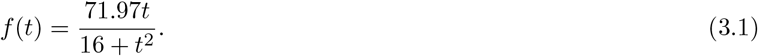

As shown in [49], taking the average of this function from average time of entering the infectious *I* class (i.e., 3 days) to the average time of recovery in this class (i.e., 8 days) gives an average fecal viral shedding rate for an infectious individual (here, individuals in compartment *I*) to be 7.65 log_10_ viral RNA per gram, which is equivalent to 4.49 *×* 10^7^ viral RNA per gram. Thus, *ζ*_*I*_ = 4.49 *×* 10^7^ viral RNA per gram. It is assumed that the average fecal viral shedding rate for hospitalized individuals (who have an average recovery period of 8 days [80]) is the same (i.e., *ζ*_*H*_ = *ζ*_*I*_ = 4.49 *×* 10^7^ viral RNA per gram). Admittedly, it is an extremely crude way of approximating *ζ*_*H*_, however since the proportion of individuals who are in *H* class is a small fraction of that in *I* [81], the impact of this crude assumption is negligible on the overall viral concentration in wastewater. Similarly, the average fecal viral shedding rate for recovered individuals who are still shedding (i.e., individuals in *J* compartment), denoted as *ζ*_*J*_, is:

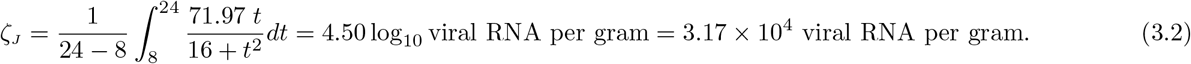

The value of the parameter *ζ*_*J*_, obtained from Equation 3.2, is significantly lower than that of *ζ*_*I*_ given in Table 3), suggesting that viral shedding recovered individuals shed at a rate that is significantly lower than that of infectious individuals. However, the actual shedding rate of the viral shedding recovered individuals may be much larger (it should be recalled that the functional form for viral shedding rate given by Equation 3.1 (derived in [49]) is based on SARS-CoV-2 hospitalized patient’s stool data in [79], and not for individuals who recovered from the infection. Thus, there is a dearth for data to use to precisely estimate the viral shedding rate by recovered individuals). Hence, conducting clinical experiments to generate the data needed to accurately quantify this parameter will be vital in improving the effectiveness of wastewater-based epidemiological modeling of the spread and control of SARS-CoV-2 in a community. The average time from excretion of feces to collection at the treatment plant is assumed to be 1 day. Thus, *d*_*V*_ = 1 *per* day (or 7 *per* week). This is a reasonable estimate, considering the fact some studies have estimated the travel time for wastewater treatment plants serving similar sized population to be between 18 to 36 hours [49, 82].

### 3.2 Estimated (fitted) parameters of the WBE model

In this section, the values of the 5 unknown parameters (also known as *estimated parameters*) of the WBE model {(2.2),(2.3)} will be obtained by fitting the model with the weekly wastewater data for the SARS-CoV-2 pandemic for the period from July 3rd, 2021 to October 9th, 2021. The model fitting is done using nonlinear least-squares optimization method [83]. In particular, MATLAB’s minimization function *lsqcurvefit* is used to minimize the sum of the squared differences between each weekly wastewater data point (data obtained from Biobot Analytics [73] and the Water and Sewer Department of Miami-Dade county [76]) and the corresponding wastewater data generated by solving the WBE model {(2.2),(2.3)}. The results obtained, for the model fitting, is depicted in Figure 3(a), showing a very good fit. Furthermore, the values of unknown parameters obtained from the data fitting are tabulated in Table 4. The projection of the number of weekly hospitalized individuals, using fixed and fitted parameters in Tables 3 and 4, respectively, is depicted with a cyan curve in Figure 3(b), alongside observed hospitalization data points marked in red dots (data obtained from The New York Times [84]). It can be seen from Figure 3 (compare (a) and (b)) that the weekly SARS-CoV-2 hospitalization data peaks a week later than the weekly wastewater data (the time when wastewater and hospitalization data peaks are marked by dashed orange and violet lines in Figures 3(a) and (b), respectively). Furthermore, Figure 3(b) shows that the model projection (cyan curve) captures the week in which weekly hospitalization peaks. It also captures the overall trend of hospitalization during the wave. The fixed and estimated parameter values, tabulated in Tables 3 and 4, respectively, were used to generate a plot for the cumulative viral RNA load, as a function of time. The results obtained, depicted in Figure 3(c), shows a good fit, as well as a very good projection of the model. Similar result is obtained for fitting the cumulative hospitalization and projection (Figure 3(d)). The plots in Figures 3(c) and (d) the WBE model (simulated using the fixed and estimated parameter values in Tables 3 and 4, respectively) can explain the cumulative data.

The parameter values obtained through data fitting are briefly discussed. The transmission rate associated with individuals in the *H* class is *β*_*H*_ = 0.00847 *per* week, which suggests that the level of SARS-CoV-2 transmission in hospitals is relatively low. The value for the parameter for the hospitalization rate (*ϕ*) is *ϕ* = 0.0066 *per* week, which suggests that only a small percentage of individuals infected with the disease during this wave in the county were actually hospitalized. Using the values of *γ*_*I*_ and *ϕ* from Tables 3 and 4 into the expression for infection-to-hospitalization ratio (IHR), given by 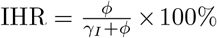 gives IHR = 0.58%, suggesting that about 0.6% of the SARS-CoV-2 infected individuals were hospitalized during the wave under consideration in this study. This result is in line with the finding in [85], which suggests, based on a statewide prevalence data from Indiana state, a range of IHR from 0.4% (for individuals younger than 40 years of age) to 9.2% (for individuals older than 60 years of age). The median age of individuals in Miami-Dade county is 41.0 years [86]. Hence, an IHR of 0.58% for Miami-Dade seems quite reasonable.

The proportion of shed viral RNA that makes it to treatment plant was estimated to be *η* = 0.96, which suggests that, in Miami-Dade, during the time period considered in this study, only a small proportion of the viral RNA shed into the wastewater was lost due to environmental factors, such as temperature-induced degradation of viral RNA. Lastly, the average fecal load *per* person *per* week was estimated to be *α* = 3283.5 grams *per* week, which is equivalent to 469.1 grams *per* day. This result is in line with the Rose *et al*. [87] study, which estimated daily fecal load to be between 51 to 796 grams *per*. Additionally, an average person of 70 kilograms (154.32 pounds) weight is estimated to excrete about 500 grams of stool *per* day [88]. The average weight of male and female residents of Miami-Dade is 188 pounds and 154 pounds, respectively [89]. Hence, assuming an equal sex ratio (i.e., an equal number of male and female in the population), the average weight of Miami-Dade residents can be estimated to be 171 pounds. Thus, an average fecal load of 469.1 grams *per* person *per* day appears to be a reasonable estimate.

The value of the basic reproduction number of the WBE model, obtained by substituting the values of the fixed and estimated parameters in Tables 3 and 4, respectively, into Equation 2.6, is *ℛ*_0_ = 2.02. Thus, since *ℛ*_0_ = 2.02 > 1, this study shows that the county was experiencing an appreciable epidemic during the duration of this particular wave.

## 4 Global Sensitivity Analysis

Global sensitivity analysis will be carried out, using Latin Hypercube Sampling (LHS) and Partial Rank Correlation Coefficients (PRCCs) [94–96], on the WBE model {(2.2)(2.3)} to determine the parameters that have the most influence on a chosen response function. Various SARS-CoV-2 modeling studies have chosen the reproduction number of the models (a measure of the average number of new cases generated by a typical infectious individual over the duration of their infectiousness) as the response function [53, 65, 97, 98]. In this study, we will use the cumulative viral load in wastewater (*W*) as the response function. Choosing the state variable *W* as the response function allows us to assess the population-level impact of the epidemiological, biological and wastewater-related parameters on wastewater itself (it is worth noting that the expression for the reproduction number of the WBE model, given by Equation (2.6), does not contain wastewater-related parameters, such as *ζ*_*I*_, *ζ*_*H*_, *ζ*_*J*_, *α, d*_*V*_ and *η*; hence, using it as a response function will not allow us to assess the impact of the wastewater-related parameters). An Increase in the rate of increase of the response function (i.e., an increase in the cumulative viral RNA concentration in the wastewater function, *W*) can signify an increase in disease burden (i.e., higher concentration of viral RNA in the wastewater means increases in the number of individuals who shed virus, and, consequently, increases in SARS-CoV-2 burden in the community). Thus, the epidemiological objective of the sensitivity analysis is to decrease the rate of increase of the response function (*W*). On the other hand, an increase in the rate of increase of the response function (*W*) can improve the precision with which wastewater surveillance can be used to accurately assess the trend, spread and burden of the disease in the community. Thus, the ecological objective is to identify the wastewater-related parameters that can improve the accuracy of the method used to measure the viral RNA concentration in the wastewater. These (epidemiological and ecological) objectives will be assessed.

The baseline values of the 13 parameters of the WBE model, given in Tables 3 and 4, will be used in the sensitivity analysis. These parameters are assumed to obey uniform distribution, and the range of each of these parameters is assumed to be 20% to the left and 20% to the right of its baseline value (except *η* which is given a range of [0.8, 1]). The range of each parameter is subdivided into 1, 000 equal sub-intervals. Since the parameter set is drawn from this set without replacement, this leads to 1, 000 *×* 13 parameter matrix (or hypercube) [53]. A parameter with a PRCC value greater than 0.5 in absolute value and corresponding small p-values (*p* < 0.05) is strongly correlated to the response function [99]. Since the response function (*W*) is a non-decreasing function, a PRCC larger than 0.5 (smaller than -0.5) indicates that a change in a parameter leads to a significant increase (decrease) in the rate of increase of the cumulative function *W*. Since the response function (*W* (*t*)) is time-dependent, the impact of the parameters of the WBE model on *W* (*t*) may vary with time. Thus, the PRCCs associated with these parameters (with respect to the response function *W* (*t*)) may be time-dependent as well.

Figure 4 depicts the time-varying PRCCs of the parameters in the response function *W* (*t*) [100] generated from the sensitivity analysis carried out in this section. This figure, which depicts the PRCC curves for parameters in the response function *W* (*t*) that had a PRCC value of higher than 0.5 in absolute value during some time *t*, shows that, while six parameters of the WBE model {(2.2),(2.3)}, namely transmission rate of infectious individuals (*β*_*I*_), average fecal load *per* person *per* unit time (*α*), average viral shedding *per* gram from individuals in *I* compartment (*ζ*_*I*_), proportion of viral RNA in the wastewater that is not lost in sewage by measurement time (*η*), progression rate from exposed to infectious class (*σ*) and rate at which excreted feces arrives at the sewage treatment plant (*d*_*V*_), are strongly positively correlated with the response function (i.e., an increase in the values of these parameters result in a corresponding increase in the rate of increase the response function), one of the parameter (recovery rate of infectious individuals (*γ*_*I*_)) is strongly negatively correlated with the response function (i.e., an increase in the value of this parameter will result in a corresponding decrease in the rate of increase of the response function). In other words, the sensitivity analysis carried out in this section identifies seven parameters of the WBE model {(2.2),(2.3)} that have the highest influence on the value of the response function, as summarized below:

**Figure 4:**
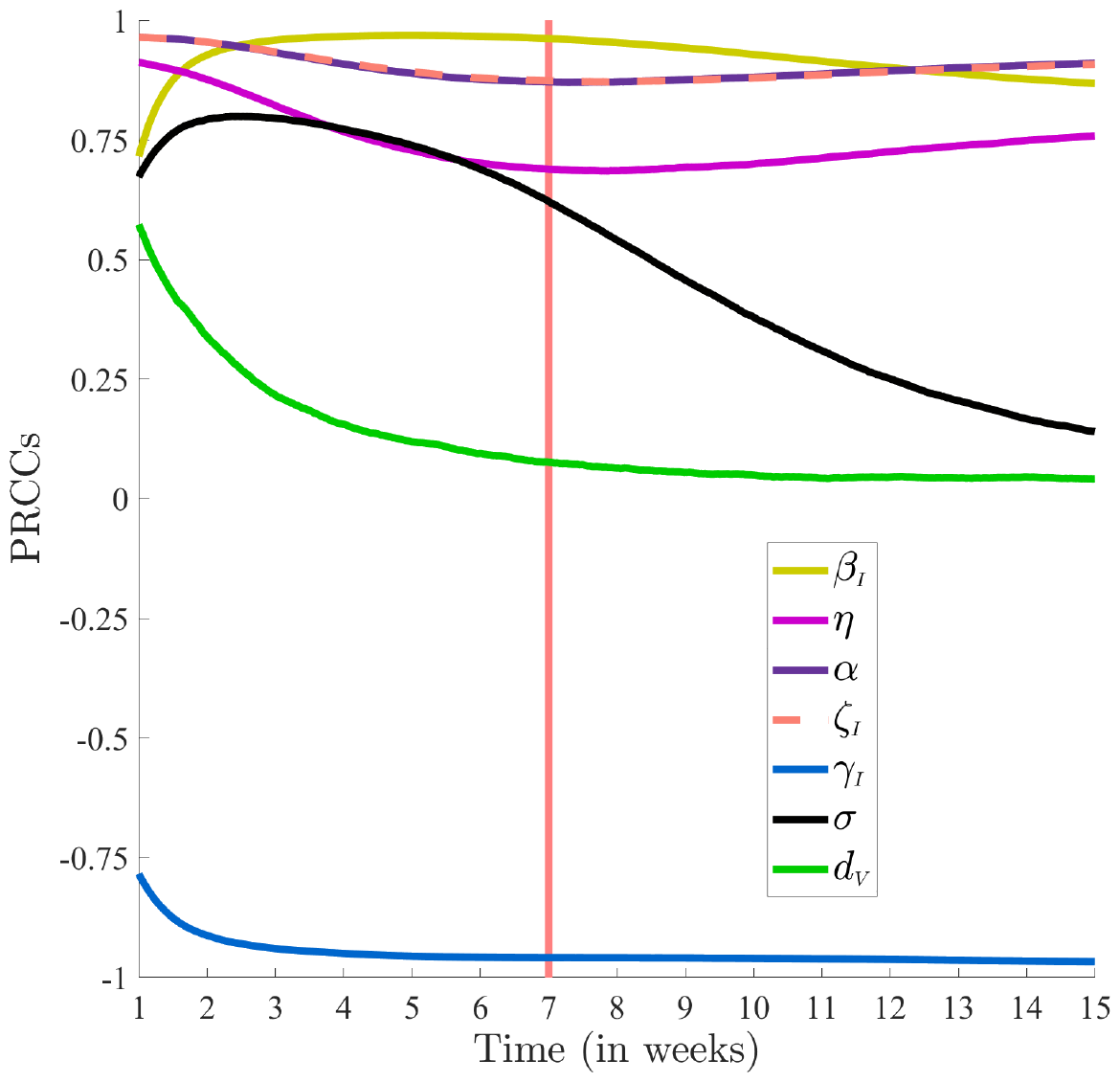
Time-varying partial rank correlation coefficients (PRCCs) of the parameters that are strongly correlated (i.e., PRCC greater than 0.5 in absolute value and corresponding small p-values (*p* < 0.05)) for the chosen response function (*W*) of the WBE model {(2.2),(2.3)} during some period of the study. Note that time-varying PRCCs of parameters *ζ*_*I*_ (depicted with a dashed orange line) and *α* (depicted with a dark violet curve) are nearly identical. Week 7 corresponds to the time when weekly wastewater data (and model projection of weekly viral load in wastewater) peaks (i.e., August 14, 2021; see Figure 3(a)) and is depicted by a vertical red line. Parameter values used in the simulations are given by the baseline values in Tables 3 and 4, and their ranges are taken to be 20% to the left and 20% to the right of the respective baseline value (except *η* which is given a range of [0.8, 1]). In this simulation p-value is less than 0.05 whenever PRCC is larger than 0.5 in magnitude.

a. The parameter for the transmission rate of infectious individuals (*β*_*I*_ ; with PRCC values lying in the range [0.72, 0.97]).
b. The parameter for the average fecal load *per* person *per* unit time (*α*; with PRCC values in the range [0.87, 0.97]).
c. The parameter for the average viral shedding *per* gram from individuals in *I* compartment (*ζ*_*I*_ ; with PRCC values in the range [0.87, 0.97]).
d. The parameter for the proportion of viral RNA in the wastewater that is not lost in sewage by measurement time (*η*; with PRCC values in the range [0.69,0.91]).
e. The parameter for the progression rate from exposed to infectious class (*σ*; with PRCC values in the range [0.14, 0.80]).
f. The parameter for the rate at which excreted feces arrives at the sewage treatment plant (*d*_*V*_ ; with PRCC values in the range [0.04, 0.57]).
g. The recovery rate of infectious individuals (*γ*_*I*_ ; with PRCCs ranging in [-0.97, -0.79]).

It can be seen from Figure 4 that although the PRCC curves for the parameters *β*_*I*_, *η, α* and *ζ*_*I*_ slightly oscillate (i.e., increase or decrease slightly) over time, their PRCC values remain consistently above +0.5. Hence, any increase in the values of these parameters will lead to a significant increase in the response function (i.e., an increase in the level of viral RNA in the wastewater) for all time *t*. For the remaining two positively-correlated parameters mentioned above (namely, *σ* and *d*_*V*_), however, Figure 4 shows that the significance of these parameters (in increasing the response function) depends on time. In particular, this figure shows that while the PRCC curves for these two parameters take values greater than +0.5 initially, the values decrease significantly to positive values much lower than +0.5 as time increases. Thus, during the first few weeks of the wave, any increase in the values of the two parameters will cause a significant increase in the response function, and an increase in the values of these parameters much later during the wave (e.g., after week 7) will only cause a marginal increase in the value of the response function. For example, the black curve in Figure 4 shows that, while during the early stages of this pandemic wave (e.g., during the first eight weeks of the wave), an increase in the value of the parameter *σ* (for the progression rate from the exposed class to the infectious class) will cause a significant increase in the viral concentration in the wastewater, an increase in the value of this parameter afterwards (i.e., after the first eight weeks of the wave) will only cause a correspondingly small increase in the viral RNA concentration in the wastewater. A similar result holds for the parameter *d*_*V*_, which is the reciprocal of the average time from excretion of feces to collection at the treatment plan.

Figure 4 shows that the only parameter that is negatively-correlated with the response function is the recovery rate parameter (*γ*_*I*_ ; with PRCC values in the range [-0.97, -0.79]). This figure (blue curve) shows a very strong negative correlation for this parameter, with the PRCC values increasing (in absolute value) from -0.79 to around -1 as time increases. Thus, this figure shows that an increase in the value of this parameter will lead to a very significant decrease in the rate of increase of the response function, *W* (*t*), for all time *t*. It is worth stating that the solid red vertical line in Figure 4 on Week 7 of the wave (corresponding to August 14, 2021) represents the time when both the weekly wastewater data (W(t)) and the model projection of weekly viral load in the wastewater peak (see Figure 3(a)). The parameters of the model that are strongly correlated with the response function during this week (when peak viral RNA is recorded by the data and predicted by the model) are:

a. The transmission rate of infectious individuals (*β*_*I*_ ; with PRCC value equals +0.96).
b. The recovery rate of infectious individuals (*γ*_*I*_ ; with PRCC value *−*0.96)
c. The average fecal load *per* person *per* unit time (*α*; with PRCC value equal + 0.86).
d. The average viral shedding *per* gram from individuals in *I* compartment (*ζ*_*I*_ ; with PRCC value equals + 0.86).
e. Proportion of shed viral RNA that is not lost in sewage by measurement time (*η*; with PRCC value + 0.69).
f. Progression rate from exposed to infectious class (*σ*; with PRCC value + 0.62).

Thus, during the week when the viral RNA peaks in the wastewater (which is also matched by the prediction of the WBE model), the top two parameters of the model that are most influential to the response function, *W*, are the transmission rate (*β*_*I*_) and the recovery rate (*γ*_*I*_). These two parameters have equal PRCC value in magnitude. The next four parameters that have the most influence on the response function are the fecal load parameter (*α*), the viral shedding parameter for infectious individuals (*ζ*_*I*_), the parameter for viral RNA not lost in sewage during measurement (*η*) and the progression rate from exposed to the infectious class (*σ*). It should be mentioned that Figure 4 only depicts the time-varying PRCC curves for parameters that have PRCC value greater or equal to 0.5 in magnitude at some time *t* during the wave (the PRCC curves for the remaining six of the WBE model that have PRCC values never exceeding 0.5 in magnitude for all time *t*, namely, *βH, ϕ, γ*_*H*_, *γ*_*J*_, *ζ*_*H*_ and *ζ*_*J*_, are depicted in Figure 5.

**Figure 5:**
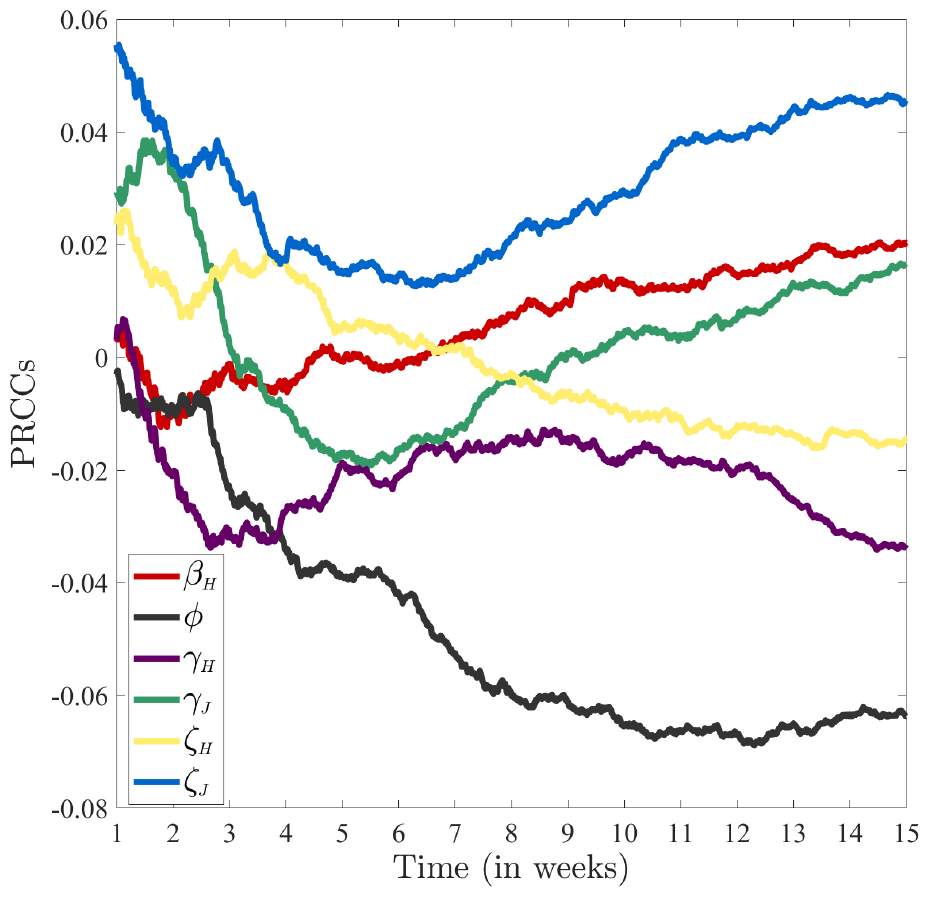
Time-varying partial rank correlation coefficients (PRCCs) of the parameters that are not strongly correlated with respect to the response function (*W*) of WBE model {(2.2),(2.3)}. Parameter values used in the simulations are given by the baseline values in Tables 3 and 4, and their ranges are taken to be 20% to the left and 20% to the right of the respective baseline value (except *η* which is given a range of [0.8, 1]).

The results of the sensitivity analysis suggest that the rate of increase of the response function (*W*) is highly correlated with increases in the transmission rate (*β*), the progression rate from exposed to infectious class (*σ*), the proportion of viral RNA that is not lost by measurement time (*η*), the rate at which excreted feces arrives at the sewage treatment plant (*d*_*V*_), the fecal load parameter (*α*), the average viral shedding rate by infectious individuals (*ζ*_*I*_) and the recovery rate (*γ*_*I*_). The epidemiological objective of the sensitivity analysis is to decrease the viral RNA in the wastewater, which entails decreasing the rate of increase of the response function *W*. Hence, it follows from the sensitivity analysis that the implementation of targeted public health control and mitigation strategies that reduce the transmission rate (*β*), the progression rate (*σ*) and increase the recovery rate (*γ*_*I*_) will significantly reduce the burden of the pandemic in the county. While the transmission rate parameter *β*_*I*_ can be reduced through nonpharmaceutical and pharmaceutical intervention measures, such as the use of high-impact face masks, social distancing, testing and isolation, progression rate parameter *σ* can be reduced by early detection and treatment of newly-infected individuals. Furthermore, the recovery rate parameter *γ*_*I*_ can be increased by using effective antiviral drugs, such as *remdesivir* [101]. The ecological objective of the sensitivity analysis is to accurately measure the viral RNA concentration in the wastewater, which entails increasing the rate of increase of the response function *W*. The results of the sensitivity analysis suggest that the implementation of strategies that increase the rate at which excreted feces arrives at the sewage treatment plant (*d*_*V*_) and the proportion of viral RNA in the wastewater that is not lost in sewage by measurement time (*η*) can increase the precision with which the spread of the disease can be captured through wastewater surveillance. The parameter for the viral RNA in the wastewater that is not lost in the sewage by measurement time (*η*) can be increased by enhancing the accuracy of the measurement of viral RNA concentration in the wastewater data (e.g., by increasing measurement frequency or selecting sampling location that is closer (relative to the location of treatment plants) to the inhabitants of the county). The fecal arrival rate parameter (*d*_*V*_) can also be increased by selecting sampling location that is closer to the inhabitants of the county. The remaining two top PRCC-ranked parameters, *α* (for the average fecal load *per* person) and *ζ*_*I*_ (for the average viral shedding rate for infectious individuals), are biological parameters that depend on the physiology of the infected individual, and there may not be specific strategies to target these processes. For instance, while having infectious individuals shed more virus into the wastewater (i.e., increase *ζ*_*I*_) will increase the accuracy of the viral RNA measurement in the wastewater, such high shedding rate increases the response function (since the parameter is highly positively-correlated with the response function), hence increasing the disease burden. In other words, more shedding increases both viral RNA measurement accuracy (a desirable feature) and disease burden (an undesirable feature). It is worth restating that while five of the seven top PRCC-ranked parameters (*β*_*I*_, *η, α, ζ*_*I*_ and *γ*_*I*_) remain highly-correlated with the response function (i.e., they have PRCC value greater or equal to 0.5 in magnitude) throughout the duration of the pandemic wave considered in this study, the remaining two top PRCC-ranked parameters (*σ* and *d*_*V*_) lose influence over time during the duration of the wave (by having PRCC values lower than 0.5 in magnitude). In summary, the sensitivity analysis conducted in this section suggests the targeted strategies that need to be implemented to ensure the effective control (or elimination) of the SARS-CoV-2 pandemic in the Miami-Dade County, as well as improving the accuracy of wastewater data in capturing the trend of the pandemic.

## 5 Numerical Simulations

In this section, the WBE model {(2.2),(2.3)} will be simulated, using the baseline values of the fixed and estimated parameters in Tables 3 and 4, to (a) predict the SARS-CoV-2 cases in Miami-Dade county, for the time period from July 3, 2021 to October 9, 2021 considered in this study and (b) quantify the contributions of recovered individuals in shedding viral RNA into the wastewater system in the county (i.e., we will quantify the contributions of viral RNA by individuals in the *J* compartment towards the weekly viral load in wastewater in the county). The simulations for the contribution of viral RNA into the wastewater by recovered individuals, which will be carried out by varying their viral shedding rate (*ζ*_*J*_) and duration of shedding (1*/γ*_*J*_), allows us to further assess the impact of two parameters of the model that were not among the most highly-correlated ones (note that this two parameters are included in Figure 5, which depicts parameters of the WBE with weaker correlations with the response function). These simulations are further described below.

### 5.1 Projecting weekly cases

The weekly SARS-CoV-2 cases predicted by the WBE model {(2.2),(2.3)} for the county will be compared with the weekly reported case data for the county, obtained from [102]. The WBE model is now simulated using the baseline values of the parameters in Tables 3 and 4, and the results obtained are depicted in Figure 6. In particular, Figure 6(a) depicts a logarithmic plot of detected SARS-CoV-2 case data for the county (magenta dots) and the model prediction of weekly cases for the county (black curve), from which it follows that a clear correlation (or consistent unimodal pattern) emerges between the data (of the weekly detected cases) and the model prediction. This figure shows an increase in the number of cases with increasing time until a peak is reached (on August 14, 2021), and the number of cases decreases thereafter. This figure also shows that the week predicted by the model for the peak number of cases in the county matches the week the peak occurred in the reported data (namely, the week of August 14, 2021; it should also be recalled from Figures 3(a) and (b) that the WBE model also accurately captured the week when the wastewater and hospitalization data peaked). It is evident from Figure 6(a) that the model projection for the weekly case data during the week of August 14 is much higher than the observed weekly case count for the county. In fact, the model prediction is about 15.37 times higher than the observed weekly case count. This result is in line with the CDC’s prediction of the true number of COVID-19-infected cases being at least 10 times higher than the reported confirmed cases [103]. Furthermore, Wu *et al*. estimate that, in 33 states in the United States (including Florida), the number of COVID-19 infections was at least 10 times larger than the number of confirmed cases [104]. We plotted the model prediction of the weekly case data for the scenario where the reported weekly case data for the county is re-scaled (i.e., multiplied) by the 15.37 factor above. The results obtained, depicted in Figure 6(b), clearly shows that the model prediction perfectly captures the trend of the reported detected weekly cases. Figure 6(c) depicts a plot of the model-predicted number of weekly cases, as a function of the detected weekly cases, showing a strong linear relationship between the two (with a correlation coefficient of *r* = 0.99 and coefficient of determination *r*^2^ = 0.98). This further confirms that the model captures the trend of the detected case data.

**Figure 6:**
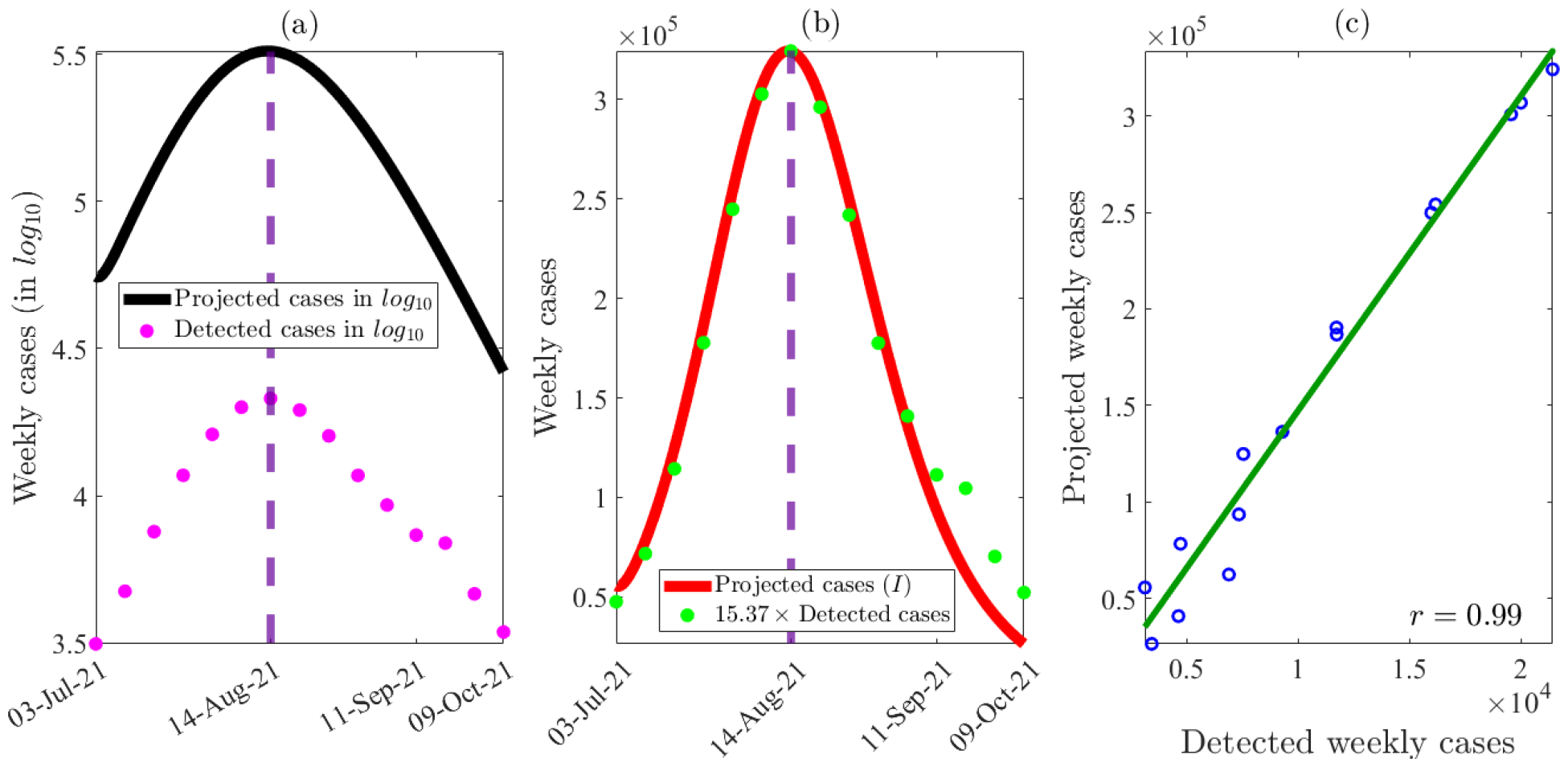
A comparison of model projected weekly cases with detected weekly cases for Miami-Dade county, Florida from July 3rd, 2021 to October 9th, 2021. (a) The log (base 10) of projected weekly cases (depicted with a black curve) along with the log of detected weekly cases obtained from Covid Act Now [102]) (depicted with red dots). (c) The model projected weekly cases (depicted with a red curve) along with 15.37-fold of the weekly detected cases (depicted with green dots). (c) The correlation of detected weekly cases with projected weekly cases (with a correlation coefficient of *r* = 0.99). The green line represents the best linear fit to the detected weekly cases versus projected weekly cases data.

### 5.2 Quantifying viral RNA contributions by recovered individuals who shed virus

The WBE model will now be simulated to quantify the contributions of viral RNA into the wastewater by individuals who newly-recovered from SARS-CoV-2 infection but are still shedding virus (i.e., individuals in the *J* class). Natarajan *et al*. [105,106] *showed that fecal SARS-CoV-2 RNA shedding can occur even seven months after infection, albeit* among a small group of individuals (3.8% of 113 patients considered in the study). Furthermore, the fecal shedding rate among individuals who have recovered but are still shedding the virus may be much higher than the baseline value used in this study (see Table 3). Hence, it is crucial to study the impact of varying the parameters related to viral shedding by recovered individuals (namely the parameters *ζ*_*J*_ and 1*/γ*_*J*_, for the shedding rate and shedding duration in the post-infectious period, respectively) on the concentration of viral RNA in the wastewater. The contribution of viral shedding at time *t*, by individuals in *K* compartment (with K ={I,H,J}), is given by:

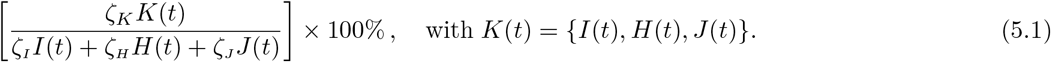

The WBE model is simulated to generate the profiles of the proportions of individuals in the infectious (*I*) and the viral shedding recovered (*J*) compartments, as a function of time. The results obtained, depicted in Figure 7(a), show that the proportions of individuals in the two viral shedding compartments increase with increasing time (i.e., as the pandemic wave progresses) until a peak is reached (which occur around August 14, 2021 for the infectious proportion and around August 28, 2021 for the recovered proportion, respectively), and decrease thereafter. This figure also shows that as the pandemic wave progresses (e.g., a month into the wave), the proportion of viral shedding recovered individuals in the community is much larger than that of the viral shedding infectious individuals. The model is further simulated to quantify the size of the population of the viral shedding recovered individuals (*J*), in relation to the population of the viral shedding infectious individuals (*I*), as a function of time (*J/I*). The results obtained, depicted in Figure 7(b), show a steady increase in this proportion with increasing time (into the pandemic wave being considered). It should be stated that, since the number of hospitalized individuals in the county is relatively small (see Figure 3(b)), we did not simulate the contribution of viral shedding by hospitalized infectious individuals.

**Figure 7:**
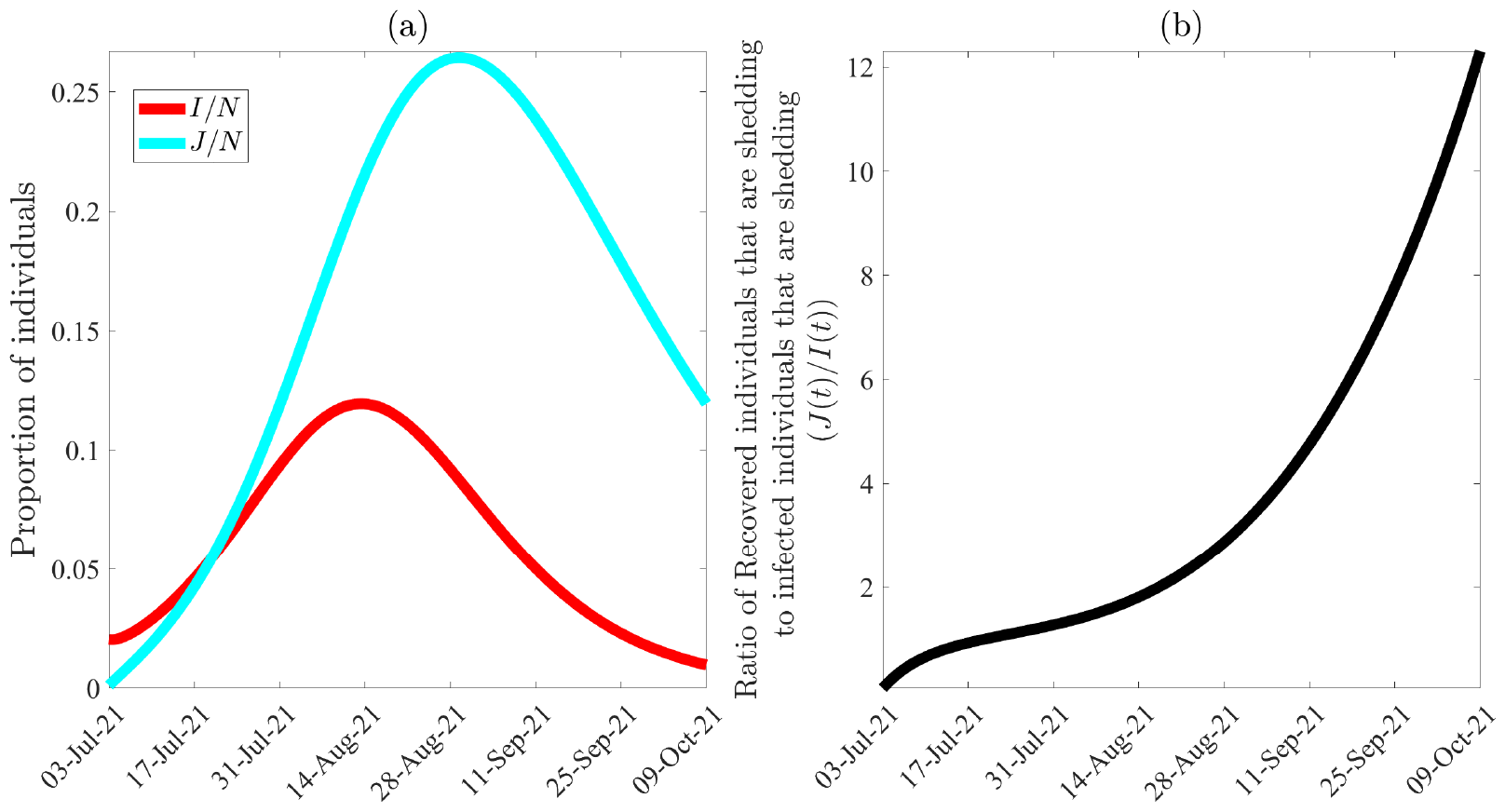
Comparison of WBE model {(2.2)(2.3)} projected profiles of the proportion of infected (*I/N*) and recovered individuals (*J/N*) who are shedding viral RNA in feces for Miami-Dade County, Florida, from July 3rd, 2021 to October 9th, 2021. (a) The profiles of the proportions of infectious (*I/N*) and recovered (*J/N*) who are shedding virus. (b) The ratio of the population of recovered individuals who are shedding to the proportion of infectious individuals (*J/I*), highlighting the relative increase in the number of recovered individuals who are shedding virus as the pandemic wave progresses. The figure is generated using the baseline values of the parameter given in Tables 3 and 4.

It should be recalled that the simulations depicted in Figure 7 show that the profile (i.e., proportion) of viral shedding recovered individuals exceeds that of viral shedding infectious individuals about a month into the wave. Yet, the viral shedding contribution of recovered individuals who shed SARS-CoV-2 into the wastewater (computed using (5.1)), under the baseline scenario (with the baseline parameter values given in Tables 3 and 4) is negligible throughout the wave. This is because, under the baseline scenario, the viral shedding rate of recovered individuals (*ζ*_*J*_) is disproportionately lower than the shedding rate of infectious individuals (*ζ*_*I*_). It is, therefore, instructive (based on the simulations in Figure 7 and that fact that the baseline value of *ζ*_*J*_ is disproportionately lower than that of *ζ*_*I*_) to explore the scenario where the viral shedding rate by recovered individuals is much higher than its baseline value To check this scenario, we simulate the WBE model to generate contour plots of the percentage of viral contribution by viral shedding recovered individuals (computed using Equation (5.1)), as a function of the parameter for viral shedding by recovered individuals (*ζ*_*J*_, but now expressed as a function of the proportion of the baseline value of the viral shedding rate of infectious individuals *ζ*_*I*_ ; that is, *ζ*_*J*_ = *cζ*_*I*_ ; where *c* ∈ [0.1, 1] and the parameter for the average shedding time by recovered individuals (1*/γ*_*J*_). The results obtained, depicted in Figure 8, show that the percentage of viral contribution by recovered individuals increases with increasing values of the viral shedding rate by recovered individuals and the average shedding time by recovered individuals. Furthermore, this figure shows that if the viral shedding rate by recovered individuals is set at one-fifth that by infectious individuals (i.e., *ζ*_*J*_ = 0.2*ζ*_*I*_), then the overall viral contribution by viral shedding recovered individuals during the second week of the wave in Miami-Dade County ranges from 8% to 12% (Figure 8(a)), depending on the average shedding duration of viral shedding recovered individuals. Similarly, for this scenario with *ζ*_*J*_ = 0.2*ζ*_*I*_, the overall viral contribution by the viral shedding recovered individuals during week seven of the wave ranges from 15% to 40% (Figure 8(b)). Thus (by comparing Figures 8 (a) and (b)), the viral contribution by viral shedding recovered individuals during week seven exceed their shedding during week two of the wave. Finally, for this scenario with *ζ*_*J*_ = 0.2*ζ*_*I*_, the simulations show that the overall viral contribution by viral shedding recovered individuals during week fifteen of the wave (i.e., at the end of the wave) ranges from 26% to 86% (Figure 8(c)). The simulation in Figure 8(c) also shows that if *ζ*_*J*_ and *γ*_*J*_ are large enough, then it is possible that the majority of the viral contribution at the end of a wave can be attributed to viral shedding recovered individuals. Hence, it can be concluded from the simulations in Figure 8 that, for the case where the viral shedding rate by recovered individuals is not disproportionately lower than that by infectious individuals the overall viral contribution by viral shedding recovered individuals during the early stage of the epidemic was low, but grows (and can dominates the contribution by infectious individuals) as the epidemic progresses through the wave. Thus, the simulations depicted in Figures 7 and 8 strongly suggest the need to conduct further lab experiments to accurately measure or estimate the viral shedding rate and shedding duration for individuals in the viral shedding recovered compartment (our study may have under-estimated the values of these two parameters; hence, further experiments to obtain more accurate values of these parameters can enhance the accuracy of the utility of wastewater-based modeling to predict the trajectory and burden of infectious diseases in a population.

**Figure 8:**
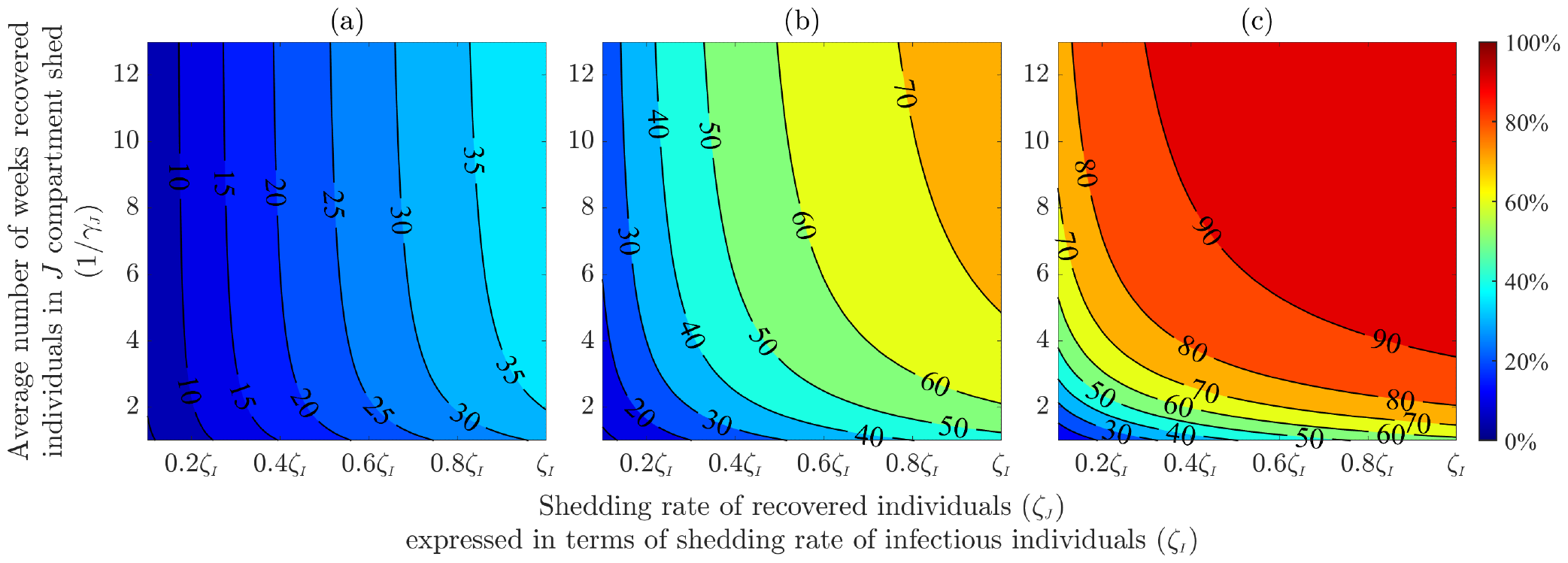
The contour plots of the percentage of viral contribution by viral shedding recovered individuals (computed using Equation (5.1)), as a function of shedding rate of viral shedding recovered individuals (*ζJ*) and the average time of viral shedding by viral shedding recovered individuals (1*/γJ*) during the (a) second week (b) seventh week and (c) fifteenth week of the study period. The x-axis of the contour plot is generated by varying *ζ*_*J*_ between 0.1*ζI* to *ζI* (where *ζI* is the viral shedding rate of infectious individuals and its value is given in Table 3) while the y-axis (i.e., 1*/γJ*) is varied from 1 to 13 weeks. The remaining parameters are taken from Tables 3 and 4.

## 6 Discussion and Conclusion

Although the use of wastewater to study the spread of infectious disease (based on isolating pathogens from the stool of some individuals infected with the disease) has a long history, perhaps dating back to the pioneering work of John Snow in the 1854s, it was only after the advent and widespread availability of efficient and safe methods for detecting viral RNA in wastewater (e.g., *reverse transcription–polymerase chain reaction*, invented in 1985 [107, 108]) that wastewater-based surveillance of disease activity has become an effective and resource-efficient tool for gathering crucial community-level public health information during outbreaks of infectious diseases [7, 8]. For example, the United States Centers for Disease Control (CDC) launched the National Wastewater Surveillance System (NWSS) in September of 2020 to “coordinate and build the nation’s capacity to track the presence of SARS-CoV-2, the virus that causes COVID-19, in wastewater samples collected across the country” [9]. Some of the benefits of wastewater-based surveillance epidemiology includes the fact that it is a less invasive surveillance mechanism (unlike the use of mass testing or large scale genomic surveillance to detect disease activity in the community) and cost-effective (in comparison to the cost of testing kits, for instance) [10, 11]. In addition to being a “leading indicator” or an “early warning signal” for disease presence or activity in a community, it can provide important insight into the presence of disease variants or their entry into community (by monitoring wastewater in airports or on airplanes) [17–21]. All in all, it is generally believed that wastewater-based surveillance epidemiology (or implementing it in combination with other forms of public health surveillance and interventions) can be a very effective tool to detect disease activity, in addition to quanttifying the trajectory and burden of the disease (thereby leading to the development of effective control and mitigation strategies) [13, 14].

Several mathematical models, of varying types (notably statistical and a few compartmental models) have been used to establish the correlation between SARS-CoV-2 RNA levels in wastewater and COVID-19 cases and burden in communities [38–47]. The current study presents a wastewater-based epidemiology (WBE) model for the dynamics of SARS-CoV-2 viral RNA in wastewater treatment plants in Miami-Dade county of Florida. The model, which takes the form of an epidemic (no human demographic) deterministic system of nonlinear differential equations (obtained by coupling equations for the dynamics of SARS-CoV-2 in humans and in the wastewater system), was fitted to weekly wastewater data for the SARS-CoV-2 pandemic in Miami-Dade for the period from July 3, 2021 to October 9, 2021. The model has a continuum of disease-free equilibria, which was shown to be locally-asymptotically stable when a certain epidemiological threshold (denoted by *R*_0_; which depends on human parameters only) is less than one. The epidemiological implication of this result is that the outbreak will not grow in the county if the reproduction threshold is brought to (and maintained at) a value less than one.

Simulations of the calibrated WBE model showed that it accurately predicts both the trend for weekly SARS-CoV-2 hospitalizations in the county and the time when th weekly hospitalizations reached a peak. Specifically, these simulations showed that there was a one-week delay between when the weekly wastewater data peaked (i.e., showed a peak in the viral RNA concentration in the wastewater; which occurred on August 14, 2021) and when the weekly hospitalization data peaked (which occurred on August 21, 2021). The ability of the WBE model to capture both peaks demonstrates its robustness to accurately predict the time delay between the two peaks. Furthermore, the calibrated WBE model was simulated to predict the weekly SARS-CoV-2 cases for the county, and the result obtained showed a very strong correlation between the predicted weekly case data and the observed weekly case data for the county. Here, too, our simulations showed that the predicted weekly case data peaked on August 14, 2021, which was exactly the same day the observed weekly case data for the county peaked. Furthermore, the maximum case count estimated by the calibrated WBE model was approximately 15 times higher than the maximum of the actual (observed or confirmed) weekly cases for the county. This result is consistent with those reported in numerous SARS-CoV-2 modeling studies (including other wastewater-based modeling, as well as statistical models) in the literature [48, 49, 104]. It is also consistent with the statement from the CDC, which estimated that their reported case count during the SARS-CoV-2 pandemic was at least 10 times lower than the actual number of infected individuals [103]. Thus, these simulations demonstrate the utility and robustness of the wastewater-based surveillance mechanism to not only detect the presence of disease in a community, but to reasonably (and, perhaps, accurately) estimate its trajectory and burden in the community.

Detailed time-varying global sensitivity analysis was carried out to determine the parameters (wastewater-based, epidemiological and biological) that have the most influence on the chosen response function (namely, the cumulative SARS-CoV-2 viral load in the wastewater system (denoted by *W* (*t*))). These analyses revealed the five parameters of the WBE model that have the highest influence on the value of the response function (*W* (*t*)) for all time *t* during the duration of the SARS-CoV-2 wave considered in this study. Specifically, the identified parameters are the transmission rate of infectious individuals (*β*_*I*_), the average fecal load *per* person *per* unit time (*α*), the average viral shedding *per* gram of infectious individuals compartment (*ζ*_*I*_), the proportion of viral RNA in the wastewater system that is not lost in the sewage by measurement time (*η*), and the recovery rate of infectious individuals (*γ*_*I*_). Thus, strategies that target these parameters (e.g., the use of face masks, testing, isolation and social-distancing that reduce the transmission rate) can reduce the overall burden of the pandemic. Furthermore, the results of the sensitivity analysis revealed that enhancing the accuracy of the measurement of viral RNA concentration in the wastewater data can be increased by increasing the proportion of shed viral RNA that is not lost in sewage by measurement time. This can be achieved by increasing measurement frequency or selecting sampling location that is closer (relative to the location of treatment plants) to the inhabitants of the county.

The simulations of the WBE model further showed that, during the period of the simulations considered, the number of recovered individuals who shed virus into the wastewater in Miami-Dade during the simulation period (i.e., individuals in the *J* compartment) exceeds that of infectious individuals (i.e., individuals in the *I* compartment) about a month into the wave. In other words, this study showed that the majority of the people shedding the SARS-CoV-2 virus into the wastewater system in Miami-Dade, during the time period considered in this study, are the viral-shedding recovered individuals (and not the viral-shedding infectious individuals). This study also showed that the viral contribution by the viral-shedding recovered individuals during the early phase of the pandemic wave considered in this study was low. That is, the majority of the viral contribution during the early stage of the outbreak may be due primarily to the infectious individuals. However, our simulations showed that the viral RNA contribution by viral-shedding recovered individuals increases with increasing time during the wave (i.e., the contribution of viral shedding recovered individuals increases as the wave progresses). In particular, the simulations showed that, during the end of the pandemic wave considered (and depending on the value of the shedding rate and the average shedding duration of viral shedding recovered individuals used in the simulations), viral-shedding recovered individuals may have accounted for the majority of the viral shedding in the Miami-Dade wastewater system. Thus, the result of this study strongly suggests the need to conduct further lab experiments to accurately measure or estimate the viral shedding rate and shedding duration for individuals in the viral shedding recovered compartment.

This study has several limitations, which we hope to account for in future studies. These include not explicitly differentiating between symptomatically-infectious individuals and asymptomatically-infectious individuals (we lumped all infectious individuals into one compartment for mathematical tractability, and owing to the fact that data to quantify viral shedding by asymptomatic infectious individuals appear to be missing at the current time). Moreover, the viral shedding rate of recovered individuals used for data-fitting purposes is significantly lower than that of infectious individuals. However, the actual rate of viral shedding of recovered individuals may be much larger. Thus, quantifying the value of this parameter based on data generated from relevant clinical (laboratory) studies will further enhance the effectiveness of wastewater-based epidemiological modeling to assess and predict the dynamics and burden of SARS-CoV-2 in a community. The study did not also include the impact of vaccination (it is intuitive to expect that viral shedding in breakthrough infections may be lower, in comparison to viral shedding by unvaccinated infected individuals; here, too, there is currently no precise data on viral shedding by vaccinated infected individuals). Furthermore, the lack of precise information about some of the dynamical aspects of the wastewater-related data also poses challenges in wastewater-based epidemiological modeling. For instance, data on average duration of fecal transportation from source to the treatment plant (i.e., data to estimate the average time from excretion of feces to arrival at the sewage treatment plant) will be very useful in the modeling process. Similarly, having daily wastewater data (rather than weekly) will help improve the accuracy of model calibration with the wastewater data (thereby improving the accuracy of the estimate of the unknown parameters from the model fitting, as well as improving the overall predictive capacity of the WBE model). Overall, this study shows that the prospect of using wastewater-based surveillance to provide insight into the transmission dynamics and effective control (and elimination) of emerging, re-emerging and resurging infectious diseases of major public health significance in a community, such as SARS-CoV-2, is very promising, particularly if such data is collected with precision and timely (e.g., daily) basis, and made readily available to modelers. We emphasize the urgent need for more clinical studies/experiments to accurately quantify the level of viral shedding by asymptomatic, recovered and vaccinated infected individuals to further enhance the predictive capacity of wastewater-based epidemiology models.

## Data Availability

Data used to calibrate the model are publicly-available (and can be shared upon request)

## Acknowledgments

ABG acknowledges the support, in part, of the National Science Foundation (Grant Number: DMS-2052363; transferred to DMS-2330801). SS acknowledges the support of the Fulbright Foreign Student Program.

### Appendix A Wastewater, Hospitalization and Case Data

**Table B.1:** Weekly wastewater, hospitalization and detected case data for Miami-Dade County, Florida from July 3, 2021, to October 9, 2021. The weekly wastewater data (given in column 2) is obtained by multiplying the average weekly processed data by Biobot Analytics [73] by 7. This weekly data is multiplied by the total influent flow rate *per* week in sewage [49], which is estimated to be 2100 million gallons *per* week for Miami-Dade County [76, 77], to generate wastewater data that is fitted with WBE model. The weekly hospitalization and case data are obtained from The New York Times [84] and Covid Act Now [102], respectively. When weekly data for a given date was unavailable, an average of the preceding and succeeding data was utilized.

| Date | Weekly wastewater data<br>(copies/mL) | Weekly hospitalization data<br>(people) | Weekly case data<br>(people) |
| --- | --- | --- | --- |
| Jul 3, 2021 | 710 | 310 | 3,138 |
| Jul 10, 2021 | 1,022 | 385 | 4,736 |
| Jul 17, 2021 | 1,610 | 537 | 7,550 |
| Jul 24, 2021 | 2,933 | 777 | 11,735 |
| Jul 31, 2021 | 4,718 | 1,120 | 16,166 |
| Aug 7, 2021 | 5,957 | 1,488 | 19,989 |
| Aug 14, 2021 | 6,041 | 1,728 | 21,415 |
| Aug 21, 2021 | 4,487 | 1,802 | 19,552 |
| Aug 28, 2021 | 4,592 | 1,788 | 15,976 |
| Sep 4, 2021 | 3,598 | 1,514 | 11,725 |
| Sep 11, 2021 | 2,457 | 1,217 | 9,301 |
| Sep 18, 2021 | 2,191 | 939 | 7,352 |
| Sep 25, 2021 | 1,442 | 778 | 6,904 |
| Oct 2, 2021 | 1,232 | 633 | 4,641 |
| Oct 9, 2021 | 917 | 512 | 3,443 |

